# Clinical and historical features associated with severe COVID-19 infection: a systematic review

**DOI:** 10.1101/2020.04.23.20076653

**Authors:** JL Pigoga, A Friedman, M Broccoli, S Hirner, AV Naidoo, S Singh, K Werner, LA Wallis

## Abstract

**Background:** There is an urgent need for rapid assessment methods to guide pathways of care for COVID-19 patients, as frontline providers need to make challenging decisions surrounding rationing of resources. This study aimed to evaluate existing literature for factors associated with COVID-19 illness severity.

**Methods:** A systematic review identified all studies published between 1/12/19 and 19/4/20 that used primary data and inferential statistics to assess associations between the outcome of interest - disease severity - and historical or clinical variables. PubMed, Scopus, Web of Science, and the WHO Database of Publications on Coronavirus Disease were searched. Data were independently extracted and cross-checked independently by two reviewers using PRISMA guidelines, after which they were descriptively analysed. Quality and risk of bias in available evidence were assessed using the Grading of Recommendations, Assessment, Development and Evaluations (GRADE) framework. This review was registered with PROSPERO, registration number CRD42020178098.

**Results:** Of the 6202 relevant articles found, 63 were eligible for inclusion; these studies analysed data from 17648 COVID-19 patients. The majority (n=57, 90·5%) were from China and nearly all (n=51, 90·5%) focussed on admitted adult patients. Patients had a median age of 52·5 years and 52·8% were male. The predictors most frequently associated with COVID-19 disease severity were age, absolute lymphocyte count, hypertension, lactate dehydrogenase (LDH), C-reactive protein (CRP), and history of any pre-existing medical condition.

**Conclusion:** This study identified multiple variables likely to be predictive of severe COVID-19 illness. Due to the novelty of SARS-CoV-2 infection, there is currently no severity prediction tool designed to, or validated for, COVID-19 illness severity. Findings may inform such a tool that can offer guidance on clinical treatment and disposition, and ultimately reduce morbidity and mortality due to the pandemic.

## INTRODUCTION

Despite containment efforts, COVID-19 has reached pandemic status. As of 21 April 2020, there are 2.5 million confirmed cases worldwide, with over 170,000 deaths;^1 2^ these numbers are only expected to grow in the coming weeks and months. Although data surrounding the novel coronavirus are rapidly evolving, initial estimates depict a dire situation: between 15% and 20% of infections lead to severe or critical disease.^2 3^ Highly vulnerable populations, compromised by factors such as malnutrition and comorbid diseases, are believed to be at greater risk of developing severe and critical disease.^4^ Mortality has varied across settings, but early data suggest a case fatality rate ranging from 0.5% to 4.5%.^5-8^ As COVID-19 spreads, healthcare systems worldwide are being strained.^9 10^ Early recognition, resuscitation and referral have proven key to effective responses, yielding lower mortality.^11^ High volumes of patients in need of critical care limit systems’ abilities to provide such care; this is already occurring even in the most highly-resourced settings.^9 10^ It is increasingly likely that the countries with the most limited capacity to respond will soon be affected on a large scale.^12^ Many low-resource settings (LRS) have scarce critical care resources, with limitations in the availability of oxygen and other basic resources as well as healthcare provider shortages.^13 14^ While ongoing acute and emergency care system strengthening efforts are underway and showing impact in many of these settings, these systems will not be able to keep pace with the emerging demand.^12^ Immediate targeted efforts are needed to assist stressed healthcare systems around the globe in managing and treating large numbers of acutely ill COVID-19 patients.^12^

Frontline providers in emergency units (EUs) need to make challenging decisions surrounding rationing of resources, including oxygen and ventilators, upon initial assessment of COVID-19 patients. Clinical guidance for evaluating patients’ respiratory needs and subsequent dispositions is essential, particularly in LRS. A range of scoring systems are available to guide care for severely ill patients. While these tools may have some utility in predicting COVID-19 patient severity, they are likely limited in this unique context. Some scores are designed for specific care settings, such as intensive care units (ICUs), a very scarce resource in LRS. ^15^ While selected COVID-19 patients can be treated in ICUs, most patients are less critical, requiring interventions more appropriate for general ward and EU settings. Thus, a tool predicting outcomes based solely on those already admitted to ICU may have reduced utility across the range of illness severity seen in this disease. Other scoring systems were purpose-designed for patients presenting with specific conditions, such as pneumonia and sepsis.^15 16^ Condition-specific tools may be of some use in assessing COVID-19 patients but are likely limited in that these patients initially present with a range of symptoms, only some of which involve respiratory disease or septic shock.^17^ High burdens of data entry seen in many tools can lead to increased time-to-decision making and frequent missing data points.^18-20^ Some also involve additional investigations, including blood tests and imaging,^15 21-23^ that have very limited availability in LRS.

In the context of the COVID-19 crisis, a low-input severity scoring tool could be a cornerstone of ensuring timely access to appropriate care and justified use of critically limited resources. In order to identify or develop a severity assessment tool appropriate for COVID-19 patients, factors contributing to, and predictive of, patient severity must first be understood. This study aimed to systematically evaluate existing COVID-19 literature for relationships between historical characteristics and clinical presentations and investigations, and illness severity.

## METHODS

A systematic review was conducted to identify papers that studied potential associations between demographics, comorbidities, clinical presentations and investigational studies, and COVID-19 illness severity. The Preferred Reporting Items for Systematic Reviews and Meta-Analysis (PRISMA) guidelines were adhered to throughout this process.^24^

### Search strategy and selection criteria

Three online databases (PubMed, Scopus, and Web of Science) were searched using a combination of free-text phrases and medical subject headings. Results were restricted to those in the English, Spanish, and Mandarin languages, published between 01 December 2019 and 19 April 2020. Search terms related to COVID-19 infection (“NCoV”, “Coronavirus”, “severe acute respiratory syndrome coronavirus 2”, “COVID”, “COVID-19”, “Coronavirus infections”) and illness severity (“Severity of Illness Index”, “severity”, “critical”, “critical illness”, “critical care”, “patient admission”, “length of stay”, “outcome”, “morbidity”, “mortality”, “death”, “respiratory insufficiency”, “respiratory distress syndrome, adult”, and “respiration, artificial”) were used in combination with those for demographics (“demographic”, “demographic”, “age”, “sex”, and “gender”), historical features (“comorbidity”, “co-morbidity”), “pre-existing”, and “condition”), clinical presentation (“signs and symptoms”, “symptom”, “characteristic”, “clinical” and, “presentation”), and diagnostic investigations (“investigation”, “laboratory”, “blood test”, “imaging”, “X-ray” and, “CT”). Refer to appendix 1 for full search strategy.

All publications in the World Health Organization’s (WHO) Database of Publications on Coronavirus Disease (COVID-19) were also included.^25^ Reference lists of all texts eligible full texts were reviewed to identify additional relevant literature.

Duplicates were manually removed. All articles were screened for inclusion by two independent reviewers (JLP, KW, SS, AVN), after which a third reviewer (AF, JLP, MB, SS, AVN) retrieved each eligible full-text and considered for inclusion. Two discrepancies in agreement were resolved through discussion.

This study included all full-text manuscripts, both pre-print and published, that provided data on associations between the outcome of interest - disease severity - and demographics, comorbidities, symptoms, vital signs, and/or investigational studies of confirmed COVID-19 patients admitted to hospitals. Research-based correspondence pieces were also included, as this is an additional format of rapid research dissemination in the COVID-19 era. Those not directly evaluating primary data, including literature reviews and meta-analyses, were excluded, as were opinion and editorial pieces, case reports, and clinical management guidelines. A range of definitions and parameters of illness severity were considered acceptable as comparator outcomes, including disease severity itself (as defined by the article’s authors), as well as proxy measures for disease severity, such as inpatient mortality and ICU admission ^26^. Manuscripts that did not include statistical analyses evaluating significance were excluded. There were no restrictions to study design. Studies evaluating exclusively children or pregnant populations were excluded.

### Data analysis

Data extracted from each article included author(s); title; countries involved; study population demographics; measure and definition of disease severity; and each presenting symptom, comorbidity, initial vital sign, and investigational study that was statistically assessed against disease severity, along with notation of whether each variable was found to be significantly associated with severity. Variables reported with a p-value ≤0.05 were considered significantly associated with illness severity.

One reviewer (AF, JLP, MB, SH, SS, AVN) extracted data, and second reviewer (AF and JLP) cross-checked all extractions. All extracted variables were documented categorically as ‘significant’, ‘not significant’ or ‘not reported’. Grading of Recommendations, Assessment, Development and Evaluations (GRADE) framework^27^ was used to assess quality of and bias in study evidence. Two reviewers (AF and JLP) graded each study, reaching consensus via discussion when necessary.

A descriptive analysis of the number of papers that reported each variable as significantly associated with illness severity was conducted. No tests of statistical significance were conducted.

### Patient and public involvement

Given the nature of this review, it was not appropriate to involve patients or the public in this study’s design or execution.

## RESULTS

The literature search identified 6202 articles across four databases (Figure 1). Thirty-one relevant publications referenced in the databases’ articles were also found. After removing duplicates, 5770 studies remained; title and abstract screening led to the removal of 5361 (92·9%) of these as they did not meet inclusion criteria. Upon review of 409 full-texts, 350 articles were excluded: 289 (82·6%) did not include significance testing for relevant variables, 55 (15·7%) did not evaluate primary data, and 2 (0·6%) were in a language other than English, Mandarin, or Spanish.

**Figure 1.**
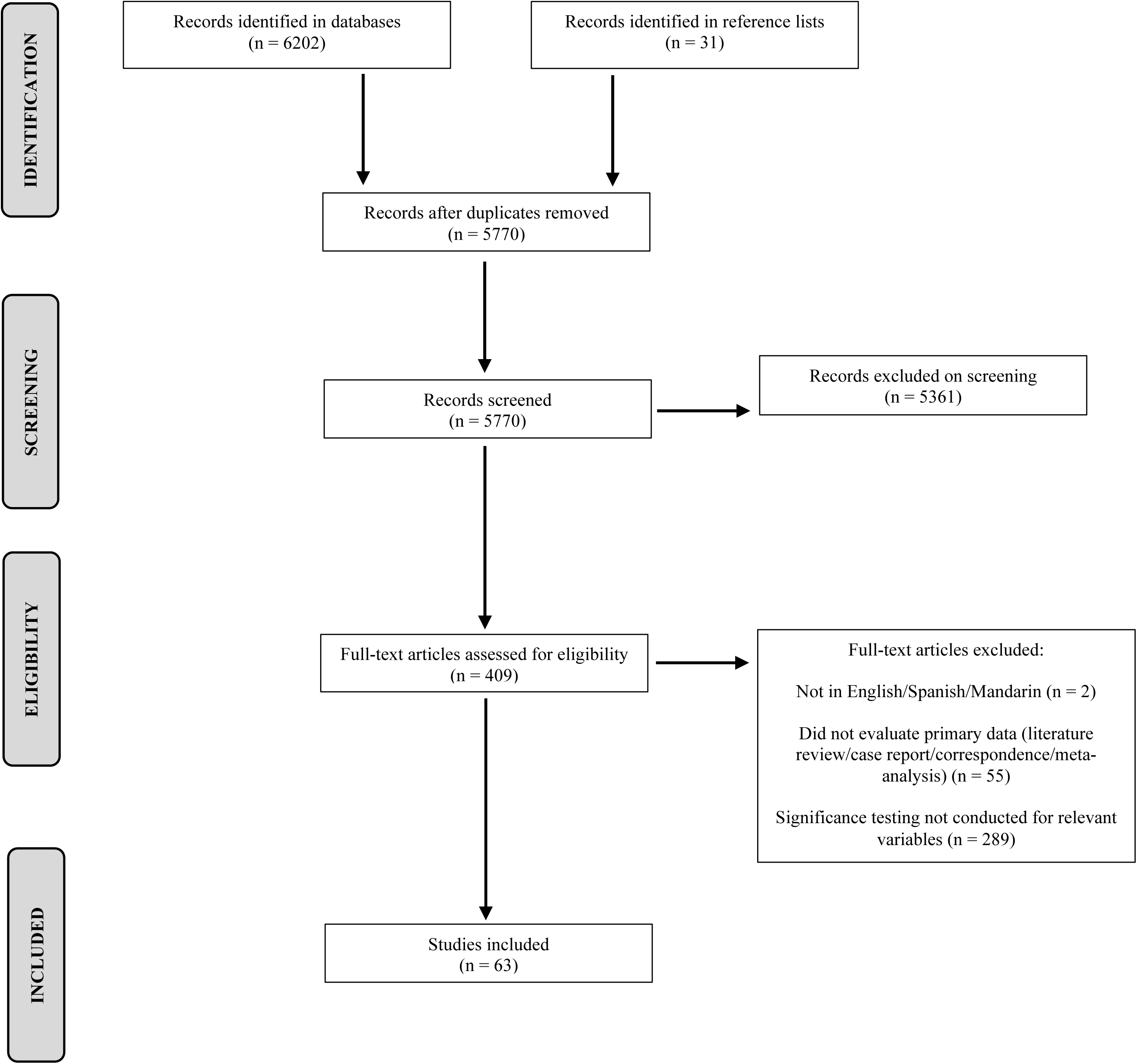
PRISMA flow diagram

Sixty-three articles were eligible for inclusion (Table 1). The majority (n=57, 90·5%) were from China; two publications were available from the United States and one each from Italy, France, South Korea, and Singapore. Most publications (n=61, 96·8%) were in English; two (3·2%) were only available in Mandarin. All studies were observational cohort studies; 57 (90·5%) retrospective and six (9·5%) prospective. Due in large part to study design, nearly all (n=62, 98·4%) studies were of low quality. One (1·6%) study was of moderate quality. Together, these studies analysed data from 17648 patients diagnosed with COVID-19 by laboratory technique (PCR or RT-PCR). Sample sizes in these studies ranged from 12 to 4103 patients, with a mean of 280. Of the 58 publications reporting sex, a median 52·8% patients were male. The median age was 52·5 years across 56 reporting studies. Eight publications included both children and adults. Four publications focused on specific populations, including healthcare workers (n=2), and those with cancer (n=1), and cardiovascular disease (n=1); the remaining 59 studies noted no population restrictions. Approximately 41% (n=26) of publications defined illness severity by the National Health Committee of the People’s Republic of China’s (NHC) guidelines (Table 2) ^25^. Fourteen studies (22·2%) evaluated inpatient mortality, and another six (9·5%) studied ICU admission.

**Table 1.** Study characteristics.

| Title | First author | Year | GRADE* | Location | Design | Population | Gender (% male) | Median age | No. severe cases | Definition of severe illness | No. variables reported on** (n) | No. variables reported as significant** (n) |
| --- | --- | --- | --- | --- | --- | --- | --- | --- | --- | --- | --- | --- |
| Clinical features of patients infected with 2019 novel coronavirus in Wuhan, China<sup>40</sup> | Huang | 2020 | Low quality | Wuhan, China | Prospective cohort | 41 adult patients with nCoV-19 admitted to a designated Wuhan Hospital from 16/12/19 - 2/1/20 | 73 | 49 | 13 | ICU admission | 42 | 16 |
| Clinical characteristics and outcomes of 112 cardiovascular disease patients infected by 2019-nCoV<sup>41</sup> | Peng | 2020 | Low quality | Wuhan, China | Retrospective cohort | 112 adult patients with nCoV-19 combined with CVD admitted to Western Hospital of Wuhan Medical College from 20/1/2020 - 15/2/2020 | 47 | 62 | 16 | NHC - critical | 30 | 6 |
| Clinical and biochemical indexes from 2019-nCoV infected patients linked to viral loads and lung injury<sup>42</sup> | Liu | 2020 | Low quality | Shenzhen, China | Retrospective cohort | 12 patients with nCoV-19 admitted to in Shenzhen Third People's Hospital through 21/1/20 | 67 | 62 | 8 | NHC - severe & critical | 8 | 8 |
| Elevated exhaustion levels and reduced functional diversity of T cells in peripheral blood may predict severe progression in COVID-19 patients<sup>43</sup> | Zheng | 2020 | Low quality | Kunming, China | Retrospective cohort | 16 patients with nCoV-19 admitted to Yunnan Provincial Hospital of ID | 56 | Not reported | 6 | No specific definition of severity provided | 7 | 5 |
| Analysis of clinical characteristics of 29 cases of new type coronavirus pneumonia<sup>44</sup> | Chen | 2020 | Low quality | Wuhan, China | Retrospective cohort | 29 patients with nCoV-19 admitted to Tongji Hospital in January 2020 | 72 | 56 | 14 | NHC - severe & critical | 9 | 2 |
| Analysis of clinical characteristics of 30 cases of new coronavirus pneumonia in medical staff<sup>45</sup> | Liu | 2020 | Low quality | Wuhan, China | Retrospective cohort | 30 adult hospital staff workers with nCoV-19 admitted to Jiangnan Hospital from 10/1/20-3/10/2020 | 50 | 35 | 4 | NHC - severe & critical | 8 | 8 |
| The clinical characteristics of myocardial injury in severe and very severe patients with 2019 Novel Coronavirus Disease<sup>46</sup> | Zhou | 2020 | Low quality | Wuhan, China | Retrospective cohort | 34 patients with nCoV-19 admitted to Tongji Medical College from 5/2/20-13/2/20 | 46 | 50 | 8 | NHC - critical | 11 | 3 |
| Diagnostic utility of clinical laboratory data determinations for patients with the severe COVID-19<sup>47</sup> | Gao | 2020 | Low quality | Fuyang, China | Retrospective cohort | 43 adult patients with nCoV-19 at Fuyang Second People's hospital from 23/1/20-2/2/20 | 60 | 44 | 15 | NHC - severe & critical | 25 | 8 |
| Quantitative computed tomography analysis for stratifying the severity of Coronavirus Disease 2019<sup>48</sup> | Shen | 2020 | Low quality | China | Retrospective cohort | 44 adult patients with nCoV-19 with CT images from 22/1/20-7/2/20 | 47 | Not reported | 9 | NHC - severe & critical | 2 | 1 |
| Relation between chest CT findings and clinical conditions of Coronavirus Disease (COVID-19) pneumonia: A multicenter study<sup>49</sup> | Zhao | 2020 | Low quality | Hunan, China | Retrospective cohort | 101 adult patients with nCoV-19 pneumonia from four institutions in Hunan | 55 | 43 | 14 | NHC - severe & critical | 4 | 2 |
| Cancer patients in SARS-CoV-2 infection: a nationwide analysis in China<sup>50</sup> | Liang | 2020 | Low quality | China | Prospective cohort | 1,590 adult patients with nCoV-19 acute respiratory disease admitted to a hospital in China | Not reported | Not reported | 131 | ICU admission, mech ventilation, mortality | 1 | 1 |
| Association of radiologic findings with mortality of patients infected with 2019 novel coronavirus in Wuhan, China<sup>51</sup> | Yuan | 2020 | Low quality | Wuhan, China | Retrospective cohort | 27 consecutive adult patients with nCoV-19 admitted to Central Hospital of Wuhan | 44 | 60 | 10 | Mortality | 13 | 6 |
| Neutrophil-to-lymphocyte ratio predicts severe illness patients with 2019 Novel Coronavirus in the early stage<sup>52</sup> | Jingyuan | 2020 | Low quality | Beijing, China | Prospective cohort | 61 adult patients with nCoV-19 infection treated at Beijing Ditan Hospital from 13/1/20-31/1/20 | 51 | 40 | 17 | NHC - severe & critical | 35 | 8 |
| Clinical features of 69 cases with Coronavirus Disease 2019 in Wuhan, China<sup>53</sup> | Wang | 2020 | Low quality | Wuhan, China | Retrospective cohort | 69 adult patients with nCoV-19 hospitalized in Union Hospital from 16/1/20-29/1/20 | 46 | 42 | 55 | Oxygen requirement <90% | 35 | 13 |
| Hematologic parameters in patients with COVID-19 infection<sup>54</sup> | Fan | 2020 | Low quality | Singapore | Retrospective cohort | 69 adult patients with nCoV-19 treated at the National Centre for Infectious Diseases in Singapore through 28/2/20 | 55 | 41 | 9 | ICU admission | 10 | 3 |
| Analysis of factors associated with disease outcomes in hospitalized patients with 2019 novel coronavirus disease<sup>55</sup> | Liu | 2020 | Low quality | Wuhan, China | Retrospective cohort | 78 adult patients with nCoV-19 admitted to three tertiary hospitals in Wuhan from 30/12/19-15/1/20 | 50 | 38 | 8 | Progression in disease state from common to severe, severe to critical or critical to death as defined by NHC | 26 | 5 |
| Clinical features and short-term outcomes of 18 patients with corona virus disease 2019 in intensive care unit<sup>56</sup> | Cao | 2020 | Low quality | Wuhan, China | Retrospective case series | 102 adult patients with nCoV-19 admitted to Wuhan University Zhongnan Hospital from 3/1/20-1/2/20 | 51 | 54 | 18 | ICU admission | 19 | 3 |
| The clinical and Chest CT features associated with severe and critical COVID-19 pneumonia<sup>57</sup> | Li | 2020 | Low quality | Chongqing, China | Retrospective cohort | 83 adult patients with nCoV-19 and abnormal findings on CT in Chongqing Medical University affiliated hospitals from 1/1/20-29/2/20 | 53 | 45.5 | 25 | NHC - severe & critical | 34 | 14 |
| Clinical predictors of mortality due to COVID-19 based on an analysis of data of 150 patients from Wuhan, China<sup>58</sup> | Ruan | 2020 | Low quality | Wuhan, China | Retrospective cohort | 150 adult patients with nCoV-19 at Jinyintan and Tongji Hospitals | 72 | 57 | 68 | Mortality | 34 | 16 |
| Clinical features and treatment of COVID-19 patients in Northeast Chongqing<sup>59</sup> | Wan | 2020 | Low quality | Chongqing, China | Retrospective cohort | 135 adult patients with nCoV-19 hospitalized in Chongqing University Three Gorges Hospital from 23/1/20-8/2/20 | 53 | 47 | 40 | NHC - severe & critical | 17 | 15 |
| Clinical characteristics of 138 hospitalized patients with 2019 novel coronavirus-infected pneumonia in Wuhan, China<sup>60</sup> | Wang | 2020 | Low quality | Wuhan, China | Retrospective case series | 138 consecutive adult patients with nCoV-19 hospitalized in Zhongnan Hospital of Wuhan University from 1/1-28/1/20 | 54.3 | 56 | 36 | ICU admission | 42 | 22 |
| Clinical characteristics of 140 patients infected by SARS-CoV-2 in Wuhan, China<sup>61</sup> | Zhang | 2020 | Low quality | Wuhan, China | Retrospective cohort | 140 adult patients with nCoV-19 infection No. 7 Hospital of Wuhan from 16/1/20-3/2/20 | 50.7 | 57 | 59 | NHC - severe | 20 | 2 |
| Clinical characteristics of refractory COVID-19 pneumonia in Wuhan, China<sup>62</sup> | Pingzheng | 2020 | Low quality | Wuhan, China | Retrospective cohort | 155 consecutive adult patients with nCoV-19 infection at Zhongnan Hospital of Wuhan University from 1/1/20-5/2/20 | 55.5 | 54 | 85 | Hospital stay <10 days | 15 | 12 |
| Abnormal coagulation parameters are associated with poor prognosis in patients with novel coronavirus pneumonia<sup>63</sup> | Tang | 2020 | Low quality | Wuhan, China | Retrospective cohort | 183 adult patients with nCoV-19 infection at Tongji Hospital 1/1/20-3/2/20 | 53.6 | 54.1 | 21 | Mortality | 9 | 5 |
| Clinical course and risk factors for mortality of adult inpatients with COVID-19 in Wuhan, China: a retrospective cohort study<sup>39</sup> | Zhou | 2020 | Low quality | Wuhan, China | Retrospective cohort | 191 patients with nCoV-19 infection at Jinyintan Hospital and Wuhan Pulmonary Hospital through 20/1/20 | 62 | 56 | 54 | Mortality | 37 | 24 |
| Risk factors associated with Acute Respiratory Distress Syndrome and death in patients with Coronavirus Disease 2019 pneumonia in Wuhan, China<sup>64</sup> | Wu | 2020 | Low quality | Wuhan, China | Retrospective cohort | 201 adult patients with nCoV-19 infection at Wuhan Jinyintan Hospital from 25/12/19-26/1/20 | 63.7 | 51 | 84 | Development of Acute Respiratory Distress Syndrome | 34 | 23 |
| Clinical progression of patients with COVID-19 in Shanghai, China<sup>30</sup> | Chen | 2020 | Low quality | Shanghai, China | Retrospective cohort | 249 adult patients with nCoV-19 at Shanghai Public Health Clinical Center from 20/1/20-6/2/20 | 50.6 | 51 | 22 | ICU admission | 13 | 1 |
| Characteristics of COVID-19 infection in Beijing<sup>65</sup> | Tian | 2020 | Low quality | Beijing, China | Retrospective cohort | 262 adult patients with nCoV-19 transferred to designated COVID hospitals by Beijing EMS from 20/1/20-10/2/20 | 48.5 | 47.5 | 46 | Dyspnea or respiratory failure | 7 | 1 |
| Dysregulation of immune response in patients with COVID-19 in Wuhan, China<sup>66</sup> | Chuan | 2020 | Low quality | Wuhan, China | Retrospective cohort | 452 adult patients with 2019-nCoV infection at Tongji Hospital from 10/1/20-12/2/20 | 52 | 58 | 286 | NHC - severe | 55 | 23 |
| Host susceptibility to severe COVID-19 and establishment of a host risk score: findings of 487 cases outside Wuhan<sup>29</sup> | Shi | 2020 | Low quality | Zhejiang Province, China | Retrospective cohort | 487 adult patients with nCoV-19 in Zhejiang Province through 11/2/20 | 53.2 | 46 | 49 | Severe, "pneumonia that can be severe and characterized by fever, cough, dyspnea, bilateral pulmonary infiltrates, and acute respiratory injury" | 8 | 6 |
| Kidney disease is associated with in-hospital death of patients with COVID-19<sup>67</sup> | Cheng | 2020 | Moderate quality | Wuhan, China | Prospective cohort | 701 adult patients with nCoV-19 infection treated at Tongji Hospital from 28/1/20-11/2/20 | 52 | 63 | 113 | Mortality | 17 | 11 |
| Anticoagulant treatment is associated with decreased mortality in severe coronavirus disease 2019 patients with coagulopathy<sup>68</sup> | Tang | 2020 | Low quality | Wuhan, China | Retrospective cohort | 449 consecutive adult patients with nCoV-19 infection treated at Tongji Hospital from 1/1/20-13/2/20 | 59.7 | 65.1 | 134 | Mortality | 7 | 6 |
| Early antiviral treatment contributes to alleviate the severity and improve the prognosis of patients with novel coronavirus disease (COVID-19)<sup>69</sup> | Wu | 2020 | Low quality | Jiangsu & Anhui Provinces, China | Retrospective cohort | 280 patients with nCoV-19 infection treated at four hospitals in the Jiangsu and Anhui from 20/1/20-19/2/20 | 53.93 | 43.12 | 83 | NHC - severe & critical | 17 | 32 |
| Clinical features of COVID-19 in elderly patients: A comparison with young and middle-aged patients<sup>28</sup> | Liu | 2020 | Low quality | Hainan, China | Retrospective cohort | 56 patients with nCoV-19 infection treated at Hainan Provincial People's Hospital from 15/1-18/2/20 | 55.4 | 53.75 | 6 | Mechanical ventilation | 1 | 1 |
| Enteric involvement in hospitalised patients with COVID-19 outside Wuhan<sup>70</sup> | Wan | 2020 | Low quality | Guangdong, Hubei, and Jiangxi Provinces, China | Retrospective cohort | 232 adult patients with nCoV-19 infection admitted to 14 Chinese hospitals from 19/1/2020 - 6/3/2020 | 55.6 | 47.5 | 61 | Vague severe | 1 | 1 |
| Prognostic value of NT-proBNP in patients with severe COVID-19<sup>71</sup> | Gao | 2020 | Low quality | Wuhan, China | Retrospective cohort | 54 consecutive adult patients with severe nCoV-19 pneumonia admitted to Hubei General Hospital (n.d.). | 44.4 | 60.4 | 102 | Mortality | 1 | 1 |
| Clinical characteristics of 161 cases of corona virus disease 2019 (COVID-19) in Changsha<sup>72</sup> | Zheng | 2020 | Low quality | Hunan, China | Retrospective cohort | 161 adult patients with nCoV-19 infection admitted to the North Hospital of Changsha first Hospital (Changsha Public Health treatment Center) from 17/1/2020 - 7/2/2020 | 49.7 | 45 | 30 | Vague severe | 29 | 12 |
| Characteristics of Liver Tests in COVID-19 Patients<sup>73</sup> | Cai | 2020 | Low quality | Shenzhen, China | Prospective cohort | 417 adult patients with nCoV-19 infection admitted to the Third People's Hospital of Shenzhen from 11/1/2020 - 21/2/2020 | 47.5 | 47 | 91 | NHC - severe & critical | 5 | 4 |
| Predictors of Mortality for Patients with COVID-19 Pneumonia Caused by SARS-CoV-2: A Prospective Cohort Study<sup>74</sup> | Du | 2020 | Low quality | Wuhan, China | Prospective cohort | 179 consecutive adult patients with nCoV-19 infection admitted to Wuhan Pulmonary Hospital from 25/12/19 - 7/3/2020 | 54.2 | 57.6 | 21 | Mortality | 38 | 18 |
| Clinical characteristics and outcomes of older patients with coronavirus disease 2019 (COVID-19) in Wuhan, China (2019): a single-centered, retrospective study<sup>75</sup> | Chen | 2020 | Low quality | Wuhan, China | Retrospective cohort | 203 older adult patients with nCoV-19 infection admitted to Zhongnan Hospital of Wuhan University from 01/1-10/02/2020 | 53.2 | 54 | 107 | NHC - severe & critical | 1 | 1 |
| COVID-19 with Different Severity: A Multi-center Study of Clinical Features<sup>76</sup> | Feng | 2020 | Low quality | Wuhan, Shanghai and Anhui Provinces, China | Retrospective cohort | 476 adult patients with nCoV-19 infection admitted to three hospitals in China between 01/1-15/02/2020 | 56.9 | 53 | 124 | NHC - severe & critical | 46 | 32 |
| The clinical course and its correlated immune status in COVID-19 pneumonia<sup>77</sup> | He | 2020 | Low quality | Wuhan, China | Retrospective cohort | 204 adult patients with nCoV-19 infection admitted to Renmin Hospital of Wuhan University between 10/1.-13/02/2020 | 38.7 | 49 | 69 | NHC - severe & critical | 45 | 22 |
| Prediction for Progression Risk in Patients with COVID-19 Pneumonia: the CALL Score<sup>78</sup> | Ji | 2020 | Low quality | Fuyang, China | Retrospective cohort | 208 consecutive adult patients with nCoV-19 infection admitted to Fuyang Second People's Hospital or Fifth Medical Center of Chinese PLA General Hospital between 20/01-22/02/2020 | 56.2 | 44 | 40 | Progression to severe COVID-19 defined as at least one of the followings, respiratory rate $\geq 30$ breaths/min, resting oxygen saturation $\leq 93\%$, PaO<sub>2</sub>/FiO<sub>2</sub> $\leq 300$ mmHg or requirement of mechanical ventilation OR worsening of lung CT findings during the observation period. | 10 | 5 |
| Risk factors for severity and mortality in adult COVID-19 inpatients in Wuhan<sup>79</sup> | Li | 2020 | Low quality | Wuhan, China | Retrospective cohort | 548 adult patients with nCoV-19 infection admitted to Tongji Hospital from 26/1 - 5/02/2020 with follow up until 03/03/2020 | 50.9 | 60 | 269 | NHC - severe & critical | 47 | 30 |
| Analysis of myocardial injury in patients with COVID-19 and association between concomitant cardiovascular diseases and severity of COVID-19<sup>80</sup> | Chen | 2020 | Low quality | Wuhan, China | Retrospective cohort | 150 adult patients with nCoV-19 infection admitted to the fever wards Tongji Hospitals from January 2019 to February 2020 | Not reported | 59 | 24 | NHC - severe & critical | 9 | 7 |
| Clinical characteristics of non-critically ill patients with novel coronavirus infection (COVID-19) in a Fangcang Hospital<sup>81</sup> | Wang | 2020 | Low quality | Wuhan, China | Retrospective cohort | 1012 non-critically ill adult patients with nCoV-19 infection admitted to Dongxihu Fangcang Hospital between 07/02-12/02/2020 | 51.8 | 50 | 100 | Deterioration of COVID-19 defined as at least one of the following: (a) symptoms persisted or worsened for $>7$ days; (b) respiratory rate $\geq 30$ or blood oxygen saturation $93\%$, at rest; (c) pulmonary imaging showed that the lesions progressed $>50\%$ within 48 hours; or (d) deterioration of co-morbidities or complications mentioned in items (f) to (i) in manuscript. | 23 | 11 |
| Risk factors associated with disease progression in a cohort of patients infected with the 2019 novel coronavirus<sup>82</sup> | Zhou | 2020 | Low quality | Nanchang , China | Retrospective cohort | 17 adult patients with nCoV-19 infection admitted to the Ninth Hospital of Nanchang between 28/01-06/02/2020 | 35.3 | 41.5 | 5 | Aggravation from admission: increased body temperature, more serious respiratory systems and enlarging CT lung lesions | 14 | 1 |
| Clinical Features and Short-term Outcomes of 102 Patients with Corona Virus Disease 2019 in Wuhan, China<sup>83</sup> | Cao | 2020 | Low quality | Wuhan, China | Retrospective cohort | 102 adult patients with nCoV-19 infection admitted to Wuhan University Zhongnan between 03/01-01/02/2020 | 51 | 54 | 17 | Mortality | 16 | 6 |
| Clinical Characteristics of 54 medical staff with COVID-19: A retrospective study in a single center in Wuhan, China<sup>84</sup> | Chu | 2020 | Low quality | Wuhan, China | Retrospective cohort | 54 adult medical staff with nCoV-19 infection from Tongji Hospital between 07/01-11/02/2020 | 66.7 | 47 | 43 | NHC - severe & critical | 7 | 2 |
| Clinical and Transmission Characteristics of Covid-19 - A Retrospective Study of 25 Cases from a Single Thoracic Surgery Department<sup>85</sup> | Li | 2020 | Low quality | Wuhan, China | Retrospective cohort | 25 adult patients with nCoV-19 infection from Tongi Hospital's Department of Thoracic Surgery, including 13 admitted patients and 12 healthcare staff, from 01/01-20/02/2020 | 76.9 | 60.2 | 9 | NHC - severe & critical | 22 | 2 |
| Data Analysis of Coronavirus CoVID-19 Epidemic in South Korea Based on Recovered and Death Cases<sup>86</sup> | Al-Rousan | 2020 | Low quality | South Korea | Retrospective cohort | 2711 patients with nCoV19 infection with data entered into the Korea Centers for Disease Control and Prevention from 20/01-29/03 | Not reported | Not reported | 9 | Mortality | 2 | 2 |
| Obesity in patients younger than 60 years is a risk factor for Covid-19 hospital admission<sup>87</sup> | Lighter | 2020 | Low quality | New York, USA | Retrospective cohort | 3615 symptomatic adult patients with nCoV-19 infection who presented to New York University hospital system from 04/03 - 04/04/2020 | Not reported | Not reported | 431 | ICU admission or invasive ventilator documentation in electronic health record | 1 | 1 |
| Factors associated with hospitalization and critical illness among 4,103 patients with COVID-19 disease in New York City<sup>88</sup> | Petrilli | 2020 | Low quality | New York, USA | Cross-sectional analysis | 4103 patients with nCoV-19 infection treated within New York University Langone Health inpatient and outpatient systems between 01/03-02/04 with follow up through 07/04. | 50.5 | 52 | 650 | ICU admission or mechanical ventilation or discharge to hospice or death | 17 | 12 |
| Suspected myocardial injury in patients with COVID-19: Evidence from front-line clinical observation in Wuhan, China<sup>89</sup> | Deng | 2020 | Low quality | Wuhan, China | Retrospective cohort | 112 adult patients with nCoV-19 infection admitted to Renmin Hospital of Wuhan University with symptom onset from 06/01-20/02/2020 | 50.9 | 65 | 67 | NHC - severe & critical | 25 | 16 |
| Clinical characteristics and outcomes of patients undergoing surgeries during the incubation period of COVID-19 infection<sup>90</sup> | Lei | 2020 | Low quality | Wuhan, China | Retrospective cohort | 34 adult patients who underwent elective surgeries and developed nCoV-19 pneumonia post-operatively with abnormal findings on chest CT at Renmin Hospital, Zhongnan Hospital, Tongji Hospital and Central Hospital from 1/1-05/02/2020 | 41.2 | 55 | 15 | ICU admission | 21 | 4 |
| Risk factors of fatal outcome in hospitalized subjects with coronavirus disease 2019 from a nationwide analysis in China<sup>91</sup> | Chen | 2020 | Low quality | China | Retrospective cohort | 1590 adult patients with nCoV-19 infection admitted to 575 hospitals throughout China from December 2019 to 31/1/2020 | Not reported | Not reported | 50 | Mortality | 7 | 5 |
| Prognostic value of myocardial injury in patients with COVID-19<sup>92</sup> | Wang | 2020 | Low quality | Wuhan, China | Retrospective cohort | 202 adult patients with nCoV-19 infection admitted to People's Hospital of Wuhan University from 31/1-05/02/2020 | 43.6 | 63 | 33 | Mortality | 21 | 11 |
| Analysis of baseline liver biochemical parameters in 324 cases with novel coronavirus pneumonia in Shanghai area<sup>93</sup> | Qian | 2020 | Low quality | Shanghai, China | Retrospective cohort | 324 adult patients with nCoV-19 pneumonia admitted to Shanghai Public Health Clinical Center from 20/1-24/2/20 | 51.5 | 51 | 26 | NHC - severe & critical | 12 | 7 |
| High prevalence of obesity in severe acute respiratory syndrome coronavirus-2 (SARS-CoV-2) requiring invasive mechanical ventilation<sup>94</sup> | Simonnet | 2020 | Low quality | Lille, France | Retrospective cohort | 124 consecutive adult patients with nCoV-19 infection admitted to Roger Salengro Hospital at "Centre Hospitalier Universitaire de Lille" between 26/2-5/04/2020 | 73 | 60 | 85 | Invasive mechanical ventilation | 5 | 3 |
| C-reactive protein correlates with CT findings and predicts severe COVID-19 early | Tan | 2020 | Low quality | Hunan, China | Retrospective cohort | 27 consecutive adult inpatients with nCoV-19 infection admitted to Hunan Provincial People's Hospital from 18/01-10/02/2020 | 41 | 48.9 | 6 | NHC - severe | 6 | 3 |
| Baseline Characteristics and Outcomes of 1591 Patients Infected With SARS-CoV-2 Admitted to ICUs of the Lombardy Region, Italy<sup>95</sup> | Grasselli | 2020 | Low quality | Lombardy, Italy | Retrospective case series | 1591 critically ill patients with nCoV-19 infection admitted from 20/2-18/03/2020 | 82 | 63 | 405 | Mortality | 2 | 2 |
| Clinical analysis on risk factors for COVID-19 patients becoming severe patients<sup>96</sup> | Ling | 2020 | Low quality | Shanghai, China | Retrospective cohort | 292 adult patients with nCoV-19 infection admitted to Shanghai Public Health Clinical Center from 20/1-10/2/2020 | 52.7 | 49.9 | 21 | NHC - severe & critical | 20 | 17 |
| Clinical Characteristics of Older Patients Infected with COVID-19: A Descriptive Study<sup>97</sup> | Niu | 2020 | Low quality | Beijing, China | Retrospective cohort | 141 patients >50yo with nCoV-19 infection transferred from general hospitals to designated hospitals by Beijing EMS from 20/1-29/2/2020 | 56.7 | Not reported | 5 | Mortality | 1 | 1 |

\* Quality and bias were assessed using the Grading of Recommendations, Assessment, Development and Evaluations (GRADE) framework.<sup>27</sup>
\*\* Study variables included were those of interest, defined as those related to patient demographics and history, and clinical presentation and investigation.

**Table 2.** Definitions of illness severity.

| Illness severity definition | Papers using definition<br>(n (%)) |
| --- | --- |
| NHC* definition of severe & critical** | 21 (33.3) |
| Inpatient mortality | 14 (22.2) |
| ICU admission | 6 (9.5) |
| NHC* definition of severe** | 3 (4.8) |
| "Severe", without additional description | 3 (4.8) |
| NHC* definition of critical** | 2 (3.2) |
| Acute respiratory distress syndrome | 1 (1.6) |
| Aggravation following admission: increased body temperature, more serious respiratory systems and enlarging CT lung lesions | 1 (1.6) |
| Deterioration of COVID-19 defined as at least one of the following: (a) symptoms persisted or worsened for >7 days; (b) respiratory rate $\geq 30$ or blood oxygen saturation 93%, at rest; (c) pulmonary imaging showed that the lesions progressed >50% within 48 hours; or (d) deterioration of co-morbidities or complications mentioned in items (f) to (i) of manuscript. | 1 (1.6) |
| Dyspnoea or respiratory failure | 1 (1.6) |
| Hospital stay >10 days | 1 (1.6) |
| ICU admission or invasive ventilator documentation in electronic health record | 1 (1.6) |
| ICU admission or mechanical ventilation or death | 1 (1.6) |
| ICU admission or mechanical ventilation or discharge to hospice or death | 1 (1.6) |
| Invasive mechanical ventilation | 1 (1.6) |
| Mechanical ventilation | 1 (1.6) |
| Progression to severe COVID-19 by NHC severe or worsening of lung CT findings during the observation period | 1 (1.6) |
| Progression to NHC definition of severe or critical condition, or death | 1 (1.6) |
| Severe pneumonia characterised by fever, cough, dyspnoea, bilateral pulmonary infiltrates, and acute respiratory injury | 1 (1.6) |
| SpO<sub>2</sub><90% | 1 (1.6) |

\*NHC = National Health Committee of People's Republic of China.
\*\*NHC stratifies illness into three categories: **mild** (slight clinical symptoms, with few or no respiratory symptoms and no pneumonia manifestations on imaging); **severe** (respiratory distress, RR ≥ 30 breaths/min, resting SpO<sub>2</sub> ≤ 93%, and PaO<sub>2</sub>/FiO<sub>2</sub> ≤ 300mmHg); and **critical** (respiratory failure, shock, and combined failure of other organs requiring mechanical ventilation and ICU-level care)<sup>98</sup>.

The most commonly analysed variables in relation to patient severity were age, sex, absolute lymphocyte count, and hypertension. The predictors most frequently associated with COVID-19 disease severity were age, absolute lymphocyte count, hypertension, lactate dehydrogenase (LDH), C-reactive protein (CRP), and history of any pre-existing medical condition.

Age was commonly identified as a significant predictor of illness severity (Table 3). The comorbidities most often reported as significant in patients with poorer outcomes were history of any pre-existing medical condition, hypertension, diabetes, and cardiovascular disease (Table 3). The presenting symptom most frequently associated with severe disease was dyspnoea or difficulty in breathing (Table 4). All other symptoms, including fever or chills, cough, sputum production, myalgias, and diarrhoea among others, were more frequently not associated with severe disease. Six of ten (60%) studies evaluating respiratory rate for initial vitals found it to be a predictor of severity, and four of five (80%) for SpO2.

**Table 3.** Demographic and historical variables significantly associated with illness severity*.

| Variable | Reported as significant*<br>(n ( %)) | Papers reporting variable (n) |
| --- | --- | --- |
| **Demographics** |  |  |
| Age | 47 (87) | 54 |
| Sex | 16 (36) | 45 |
| **History of comorbidities** |  |  |
| Hypertension | 22 (67) | 33 |
| Any pre-existing condition | 17 (74) | 23 |
| Cardiovascular disease | 16 (64) | 25 |
| Diabetes | 13 (41) | 32 |
| Body mass index (BMI) | 6 (55) | 11 |
| Chronic obstructive pulmonary disease (COPD) | 6 (32) | 19 |
| Stroke | 5 (42) | 12 |
| Malignancy | 5 (26) | 19 |
| Chronic kidney disease (CKD) | 4 (29) | 14 |
| Other respiratory disease | 2 (29) | 7 |
| Current smoker | 1 (9) | 11 |
| Tuberculosis | 0 (0) | 3 |
| Hepatitis/cirrhosis | 0 (0) | 14 |
| Immunocompromised | 0 (0) | 2 |

\*Significance denoted by $p < 0.05$ as related to illness severity by definition specified in paper.

**Table 4.**
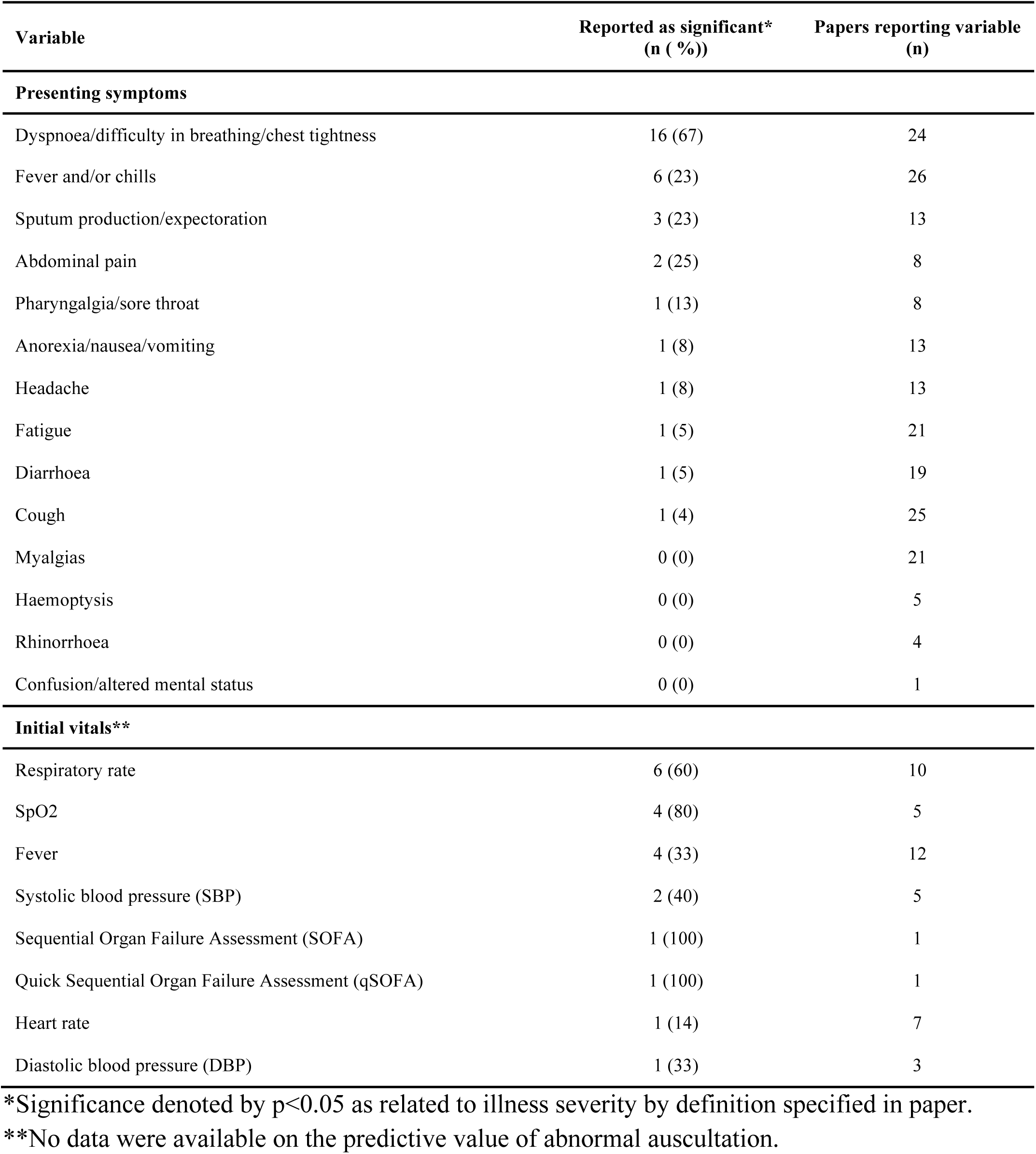
Clinical presentation variables significantly associated with illness severity*.

The most commonly associated laboratory values were absolute lymphocyte count, LDH, CRP, D-dimer, and procalcitonin (Table 5). Among associated laboratory values, PaO2/FiO2,N-terminal b-type natriuretic peptide (NT-proBNP) and blood glucose were each consistently reported as significant across multiple studies. Abnormal CT was significant in ten of 16 (62·5%) studies, and abnormal chest X-ray in two of four (50%) (Table 5).

**Table 5.**
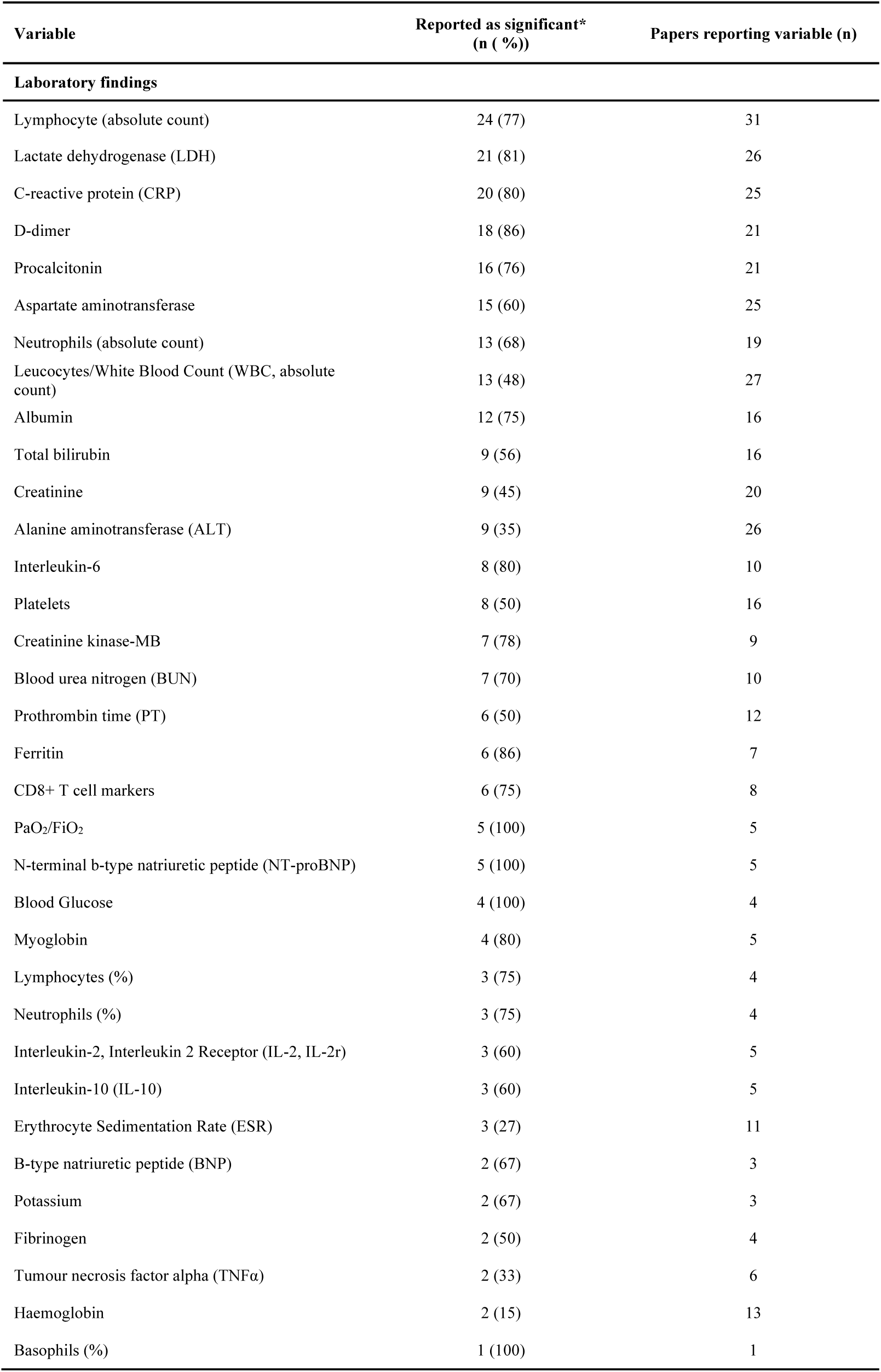
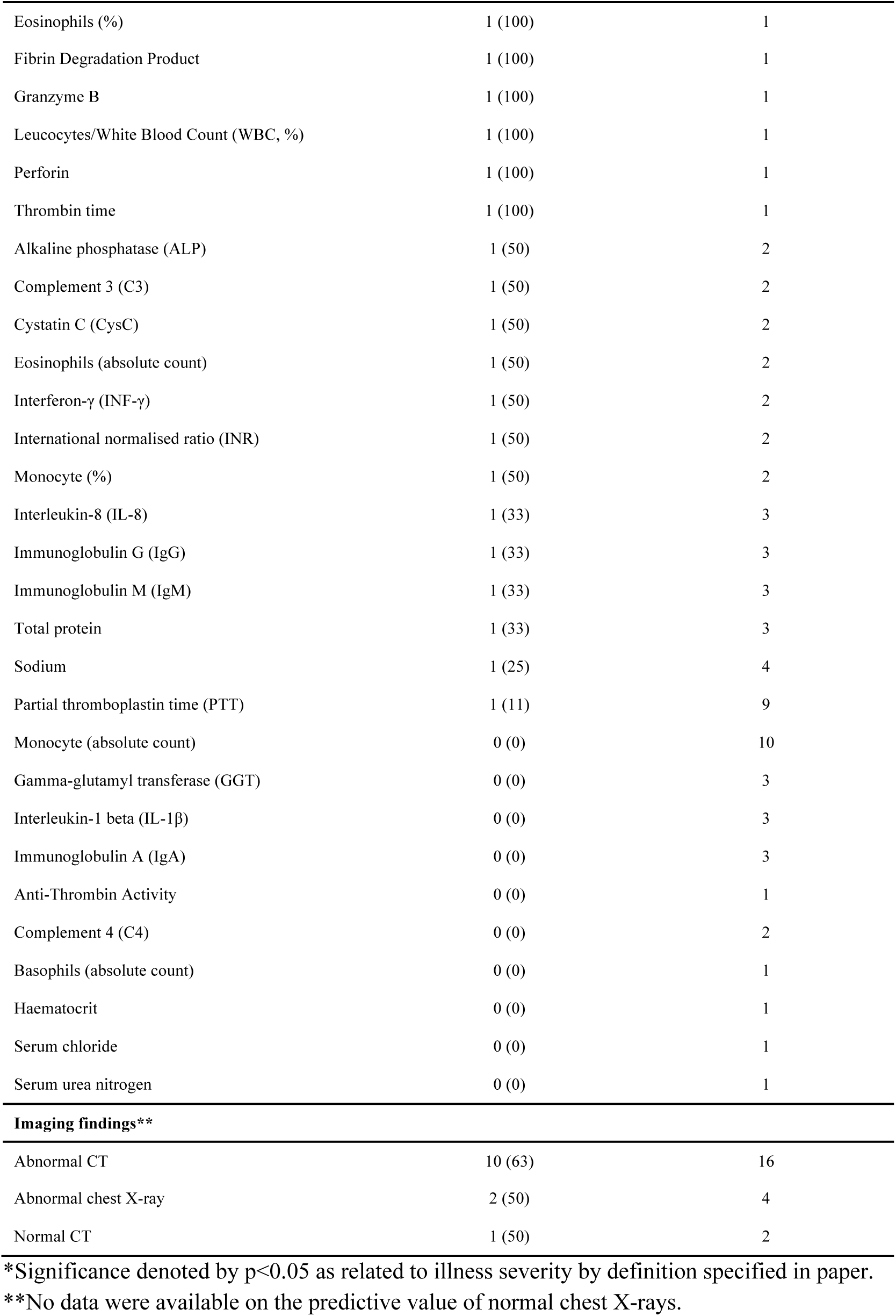
Clinical investigation variables significantly associated with illness severity*.

| Variable | Reported as significant*<br>(n (%)) | Papers reporting variable (n) |
| --- | --- | --- |
| **Laboratory findings** |  |  |
| Lymphocyte (absolute count) | 24 (77) | 31 |
| Lactate dehydrogenase (LDH) | 21 (81) | 26 |
| C-reactive protein (CRP) | 20 (80) | 25 |
| D-dimer | 18 (86) | 21 |
| Procalcitonin | 16 (76) | 21 |
| Aspartate aminotransferase | 15 (60) | 25 |
| Neutrophils (absolute count) | 13 (68) | 19 |
| Leucocytes/White Blood Count (WBC, absolute count) | 13 (48) | 27 |
| Albumin | 12 (75) | 16 |
| Total bilirubin | 9 (56) | 16 |
| Creatinine | 9 (45) | 20 |
| Alanine aminotransferase (ALT) | 9 (35) | 26 |
| Interleukin-6 | 8 (80) | 10 |
| Platelets | 8 (50) | 16 |
| Creatinine kinase-MB | 7 (78) | 9 |
| Blood urea nitrogen (BUN) | 7 (70) | 10 |
| Prothrombin time (PT) | 6 (50) | 12 |
| Ferritin | 6 (86) | 7 |
| CD8+ T cell markers | 6 (75) | 8 |
| PaO<sub>2</sub>/FiO<sub>2</sub> | 5 (100) | 5 |
| N-terminal b-type natriuretic peptide (NT-proBNP) | 5 (100) | 5 |
| Blood Glucose | 4 (100) | 4 |
| Myoglobin | 4 (80) | 5 |
| Lymphocytes (%) | 3 (75) | 4 |
| Neutrophils (%) | 3 (75) | 4 |
| Interleukin-2, Interleukin 2 Receptor (IL-2, IL-2r) | 3 (60) | 5 |
| Interleukin-10 (IL-10) | 3 (60) | 5 |
| Erythrocyte Sedimentation Rate (ESR) | 3 (27) | 11 |
| B-type natriuretic peptide (BNP) | 2 (67) | 3 |
| Potassium | 2 (67) | 3 |
| Fibrinogen | 2 (50) | 4 |
| Tumour necrosis factor alpha (TNFα) | 2 (33) | 6 |
| Haemoglobin | 2 (15) | 13 |
| Basophils (%) | 1 (100) | 1 |
| Eosinophils (%) | 1 (100) | 1 |
| Fibrin Degradation Product | 1 (100) | 1 |
| Granzyme B | 1 (100) | 1 |
| Leucocytes/White Blood Count (WBC, %) | 1 (100) | 1 |
| Perforin | 1 (100) | 1 |
| Thrombin time | 1 (100) | 1 |
| Alkaline phosphatase (ALP) | 1 (50) | 2 |
| Complement 3 (C3) | 1 (50) | 2 |
| Cystatin C (CysC) | 1 (50) | 2 |
| Eosinophils (absolute count) | 1 (50) | 2 |
| Interferon-γ (INF-γ) | 1 (50) | 2 |
| International normalised ratio (INR) | 1 (50) | 2 |
| Monocyte (%) | 1 (50) | 2 |
| Interleukin-8 (IL-8) | 1 (33) | 3 |
| Immunoglobulin G (IgG) | 1 (33) | 3 |
| Immunoglobulin M (IgM) | 1 (33) | 3 |
| Total protein | 1 (33) | 3 |
| Sodium | 1 (25) | 4 |
| Partial thromboplastin time (PTT) | 1 (11) | 9 |
| Monocyte (absolute count) | 0 (0) | 10 |
| Gamma-glutamyl transferase (GGT) | 0 (0) | 3 |
| Interleukin-1 beta (IL-1β) | 0 (0) | 3 |
| Immunoglobulin A (IgA) | 0 (0) | 3 |
| Anti-Thrombin Activity | 0 (0) | 1 |
| Complement 4 (C4) | 0 (0) | 2 |
| Basophils (absolute count) | 0 (0) | 1 |
| Haematocrit | 0 (0) | 1 |
| Serum chloride | 0 (0) | 1 |
| Serum urea nitrogen | 0 (0) | 1 |
| **Imaging findings\*\*** |  |  |
| Abnormal CT | 10 (63) | 16 |
| Abnormal chest X-ray | 2 (50) | 4 |
| Normal CT | 1 (50) | 2 |

\*Significance denoted by $p < 0.05$ as related to illness severity by definition specified in paper.
\*\*No data were available on the predictive value of normal chest X-rays.

## CONCLUSION

The novelty of SARS-CoV-2 infection has led to many hypotheses regarding potentially significant predictors of patient illness. This study aimed to provide clarity around potential predictors by presenting a systematic review of the features most frequently and consistently associated with COVID-19 illness severity. The available literature evaluates a broad range of clinical characteristics and investigations, highlighting variables likely to be significantly associated with illness severity, including age, history of pre-existing medical conditions, dyspnoea on presentation, and a range of blood tests.

In line with several publications,^28-30^ age was identified as a predictor of illness severity in nearly all studies. However, not all studies addressed confounding in their methodology. It is not clear whether age is an independent predictor for severity due to decreased physiological reserve or if increased rates of comorbidities amongst elderly populations influence its significance. Results suggest that any pre-existing condition may put COVID-19 patients at higher risk of poor outcomes. But, when looking at specific conditions, few comorbidities were found to be consistently significant across the literature. Hypertension, cardiovascular disease, and body mass index (BMI) were found to be significant predictors in the majority of instances where they were evaluated; these should be assessed further in future investigations

In clinical practice, clinical discriminators and vital signs play important roles in determining acuity. Shortness of breath or dyspnoea was the only presenting symptom reported as a significant predictor of severity across these studies. An earlier meta-analysis of COVID-19 data also supports this conclusion.^31^ Dyspnoea is easy to assess and already being used in many settings for COVID-19 screening,^31-33^ making it a candidate for potential inclusion in a COVID-19 severity scoring tool. Data on initial vital signs were inconclusive.

Although a large portion of potentially predictive variables depend on blood investigations, these predictors may have limited utility in the initial management of COVID-19 disease, especially in LRS: Faced with scarce time and resources, and overwhelming patient numbers, clinicians around the world are in need of near-immediate indicators of illness severity. Potentially relevant severity calculators such as the Pneumonia Severity Index and CURB-65, require blood investigations and imaging to predict mortality, and are based on non-COVID-19 pneumonia,^23 34^ limiting their utility in the pandemic.

Due to the novelty of SARS-CoV-2 infection, there are currently no scoring systems validated for COVID-19 illness severity assessment either at the initial point of entry to the healthcare system or during admission. Such a tool could enable rapid classification of patients’ illness severity to determine disposition and resource allocation, aiding overwhelmed healthcare systems facing limited resources. It could assist frontline providers in decision-making surrounding resource allocation, including both the physical - e.g. oxygen and mechanical ventilation - and human resources necessary to oversee such interventions in a timely, appropriate manner for the rapidly increasing volume of COVID-19 patients.

Most LRS already suffered from scarcities of oxygen, mechanical ventilation and ICU beds pre-COVID-19,^35^ and the current resource constraints seen in highly-resourced settings will likely be even more extreme in LRS.^36 37^ Ethical allocation of resources is expected to become a necessity in all settings, as providers will have to divide resources across both COVID-19 and non-COVID-19 patients, many of whom will have similar prognoses.^38^ Though a severity scoring tool may identify a patient’s need for oxygen or mechanical ventilation, broader resource allocation for all patient populations requires ethical frameworks beyond such a tool’s scope. This review’s findings may have utility as components of a COVID-19-specific severity scoring tool that could offer guidance on clinical treatment and disposition, and ultimately reduce morbidity and mortality due to the COVID-19 pandemic.

Some discrepancies exist between our findings and calculators currently being used to evaluate COVID-19 patients, namely on the significance of initial vitals, presenting symptoms, laboratory values and imaging findings. Though our findings were inconclusive in regard to initial vitals, one study reported that qSOFA was predictive of COVID-19 illness severity,^39^ indicating that altered mental status, tachypnoea and systolic blood pressure ≤100mmHg could be predictive in a severity scoring tool. Most pneumonia clinical calculators to predict illness severity rely on laboratory and imaging investigations that were insignificant in our systematic review—aside from glucose, absolute lymphocyte count and PaO2/FiO2—and are not immediately available in most LRS. Various calculators also emphasise sex and pulmonary co-morbidities, which were found to be independently insignificant in our review. The discrepancies between our systematic review and the clinical calculators of illness severity being used in this pandemic warrant a COVID-19 specific-calculator based on the emerging evidence rather than single-institution experience to facilitate illness severity stratification.

Due to the emergent COVID-19 pandemic and need for data to inform responses, rapid efforts are being made to publish any available information. Our review included research-based correspondence and pre-print articles alongside peer-reviewed publications, all of which were in the form of observational cohort studies. However, all studies performed well against GRADE criteria, scoring at least as well as is expected for cohort studies (low quality or above).

An overwhelming majority of studies originated in China; this suggests that there may be increased homogeneity in the data and that results might not be translatable to other settings. It could not be confirmed that none of the publications used populations from the same patient database.

Findings of this study may also be limited in that they are solely descriptive, lacking any testing of statistical significance. Numerous definitions for, and proxy measures of, illness severity were presented in the primary literature included in this review. While this variance may have influenced results, it could not be avoided given the urgency of the COVID-19 pandemic and lack of existing evidence.

This systematic review presents the features most associated with COVID-19 illness severity based on frequency and consistency of reporting publications. These findings could aid frontline providers in determining how to allocate scarce resources, namely oxygen and mechanical ventilation, in a timely and appropriate manner for the rapidly increasing volume of COVID-19 patients around the world, but especially in LRS. It is our hope that by using significant variables such as age, history of comorbidity, and dyspnoea as a presenting symptom, providers in LRS can quickly predict illness severity to guide clinical treatment and disposition of patients with COVID-19 disease. Given the urgency of the pandemic and the almost universally low-quality of data, future research collaborations with larger samples and prospective designs are critical to improving COVID-19 illness severity prediction and subsequent management.

## Data Availability

Additional extracted data not provided in-text or as supplementary information are available upon request.

## Contributors

LAW, JLP, and MB designed the study. JLP performed the database searches. JLP, KW, SS, and AVN screened articles titles and abstracts for inclusion. AF, JLP, MB, SS and AVN screened eligible full texts for inclusion. AF, JLP, MB, SH, SS, and AVN extracted data. JLP and AF cross-checked all data extractions, assessed quality of included studies, performed data analysis, and drafted the manuscript. All authors contributed to revising the manuscript and have approved of the final version. LAW is the guarantor.

## Appendix Search strategy

**PubMed**

Search date: 19/4/20

Restricted to studies published from 1/12/19-19/4/20

| # | Search terms | No. results |
| --- | --- | --- |
| 1 | NCoV [TIAB] OR Coronavirus [MeSH] OR "severe acute respiratory syndrome coronavirus 2" [MeSH] OR "COVID*" [TIAB] OR "COVID-19" [TIAB] OR Coronavirus infections [MeSH] or "coronavirus" [TIAB] | 5 581 |
| 2 | Severity of Illness Index [MeSH] OR "sever*" [TIAB] OR "critical*" [TIAB] OR Patient Admission [MeSH] OR Length of Stay [MeSH] OR Critical Illness [MeSH] OR Critical care [MeSH] OR Outcome* [TIAB] OR Mortality [MeSH] OR Death [MeSH] OR Morbidity [MeSH] OR Respiratory Insufficiency [MeSH] OR Respiratory Distress Syndrome, Adult [MeSH] OR Respiration, Artificial [MeSH] OR "ventilation" [TIAB] | 106 821 |
| 3 | Demography [MeSH] OR "Age" [TIAB] OR "Sex" [TIAB] OR "Gender" [TIAB] | 84 335 |
| 4 | Comorbidity [MeSH] OR "Co-morbid*" [TIAB] OR "Comorbid*" [TIAB] OR "Pre-exist*" [TIAB] OR "Condition*" [TIAB] | 18 804 |
| 5 | Signs and Symptoms [MeSH] OR "symptom*" [TIAB] OR "characteristic*" OR "clinical" OR "presentation*" [TIAB] | 20 966 |
| 6 | Diagnostic Techniques and Procedures [MeSH] OR "Imag*" OR "X-ray*" OR "Xray*" OR "CT" OR "Computed Tomography" OR "Radiograph" | 43 618 |
| 7 | 1 + 2 + 3 | 168 |
| 8 | 1 + 2 + 4 | 117 |
| 9 | 1 + 2 + 5 | 378 |
| 10 | 1 + 2 + 6 | 123 |

**Scopus**

Search date: 19/4/20

Restricted to studies published from 1/1/19-19/4/20

| # | Search terms | No. results |
| --- | --- | --- |
| 1 | TITLE-ABS-KEY(NCoV OR severe acute respiratory syndrome coronavirus 2 OR COVID* OR COVID-19 OR coronavirus) | 828 |
| 2 | TITLE-ABS-KEY("severity of illness index" OR sever* OR critical* OR "patient admission" OR "length of stay" OR "critical illness" OR "critical care" OR outcome* OR mortality OR death OR morbidity OR respiratory AND insufficiency OR "respiratory distress syndrome" OR respiration OR ventilation) | 24 468 |
| 3 | TITLE-ABS-KEY(demograph* OR age OR sex OR gender) | 132 302 |
| 4 | TITLE-ABS-KEY(comorbidity OR co-morbid* OR comorbid* OR pre-exist* OR preexist OR condition*) | 556 058 |
| 5 | TITLE-ABS-KEY("signs and symptoms" OR symptom* OR characteristic* OR clinical OR presentation*) | 1 001 095 |
| 6 | TITLE-ABS-KEY(diagnostic OR techniques OR procedures OR invest*OR imag* OR X-ray* OR Xray* OR CT OR Computed Tomography OR radiograph*) | 46 425 |
| 7 | 1 + 2 + 3 | 28 |
| 8 | 1 + 2 + 4 | 32 |
| 9 | 1 + 2 + 5 | 60 |
| 10 | 1 + 2 + 6 | 7 |

**Web of Science**

Search date: 19/4/20

Restricted to studies published from 1/1/19-31/3/20

| # | Search terms | No. results |
| --- | --- | --- |
| 1 | TS=(COVID* OR NCoV OR Coronavirus* OR SARS-CoV-2) | 1 132 |
| 2 | TS=(Sever* OR Critical* OR Admit* OR Admission* OR Length of stay* OR Critical Care OR Intensive Care OR Outcome OR Mortality OR Death OR Morbidity OR Respiratory Failure OR Respiratory Distress OR Respiration OR Ventilation) | 625 363 |
| 3 | TS=(Demograph* OR Age* OR Gender* OR Sex*) | 397 718 |
| 4 | TS=(Comorbid* OR Co-morbid* OR Preexist* OR Pre-exist* OR Condition*) | 358 857 |
| 5 | TS=(Symptom* OR Sign* OR Characteristic* OR Clinical* OR Present*) | 1 363 307 |
| 6 | TS=(Vital* OR Vital* Sign*) | 24 207 |
| 7 | TS=(Blood* OR Blood Test* OR Laboratory* OR Investigat* OR Blood Count*) | 653 308 |
| 8 | TS=(Imaging OR Image OR X-ray* or Xray* OR CT OR Computed Tomography OR Radiograph*) | 260 291 |
| 9 | 1 + 2 + 3 | 165 |
| 10 | 1 + 2 + 4 | 59 |
| 11 | 1 + 2 + 5 | 442 |
| 12 | 1 + 2 + 6 | 12 |
| 13 | 1 + 2 + 7 | 203 |
| 14 | 1 + 2 + 8 | 57 |

